# Increasing Value in the Veterans Affairs Healthcare System (VA) with Precision Health: A Continuing Landmark Collaboration with the Department of Energy

**DOI:** 10.1101/2025.08.21.25334155

**Authors:** Amy C. Justice, Benjamin McMahon, Daniel A. Jacobson, Kelly Cho, Anuj J. Kapadia, Samuel Aguayo, Zeynep H. Gümüş, Ioana Danciu, Jean C. Beckham, Nathan A. Kimbrel, Silvia Crivelli, Eilis Boudreau, Pat Finley, Alex Bryant, Michael Green, Shinjae Yoo, Jacob Joseph, Peter Reaven, Jin Zhou, Shiuh-Wen Luoh, Ravi Madduri, Ayman Fanous, Khushbu Agarwal, Harshini Mukundan, Sumitra Muralidhar

## Abstract

**Objective:** By personalizing healthcare to an individual’s specific requirements, precision health promises to maximize benefit and minimize harm, thereby maximizing value. We describe here, how in Phase 2 of the Million Veteran Program–Computational Health Analytics for Medical Precision to Improve Outcomes Now (MVP-CHAMPION), artificial intelligence (AI) and high performance computing (HPC) have been applied to Veteran’s electronic health records (EHRs) and genetic data to advance real-world precision health.

**Materials and Methods:** Eight concept projects were selected on the basis of potential impact on high-burden conditions among Veterans, including heart failure, suicide, lung cancer, diabetes, post COVID-19 sequelae, medication toxicity, and obstructive sleep apnea.

**Results:** Achievements include new and more discriminating risk prediction models to inform medical decision making, multimorbidity-aware analytic frameworks, and development and deployment of reusable computational tools. The identification of novel risk factors from genetic data and unstructured text in the EHR has both informed risk prediction and offered new insights for medication repurposing and development.

**Discussion:** We not only confirmed the need for shared infrastructure, data management, and novel AI-based workflows to inform precision health, but also found that such programmatic improvements result in valuable mechanistic insights.

**Conclusion:** By building on these foundations through expanded deployment, adaptive modeling, and broader partnerships, the VA-DOE collaboration is poised to transform not only the future of Veterans’ healthcare, but the broader national landscape of precision health.

## BACKGROUND AND SIGNIFICANCE

Among the world’s largest and wealthiest countries, the United States (US) far outspends others on healthcare and leads the world in biomedical innovation. Yet we experience the lowest life expectancy^1^ and often the healthcare we deliver fails to improve health. In short, the US faces a crisis in return on investment, or value, in healthcare. Much of our specialty medical and psychiatric care is ill suited to address the multimorbidity that becomes common as people age. Our burden of chronic, co-occuring (multimorbid), medical and psychiatric disease and rates of suicide and drug overdose exceed that of our economic peers.^1^ Rather than a “one size fits all” with a given medical diagnosis, precision or personalized health promises to improve healthcare by integrating real-world clinical, genomic, and social and environmental determinants of health data with modern computational methods to tailor or personalize care to the individual. By better differentiating those most and least likely to benefit from an intervention, personalized health could substantially improve value. However, before the benefits of personalized health can be fully realized, data scientists need to address important limitations associated with reproducibility, reliability and usability of healthcare models.

In 2016, the United States Department of Veterans Affairs (VA) and the Department of Energy (DoE) recognized the profound synergy of combining forces to address this crisis and launched MVP-CHAMPION (Million Veteran Program–Computational Health Analytics for Medical Precision to Improve Outcomes Now). VA offers a unique laboratory in which to address this crisis as it implemented a national, paperless, electronic health record (EHR) thirty years ago. Since then over 25 million Veterans have had their longitudinal health care documented in the system and over a million Veterans have provided genetic data linked to their EHR data by enrolling in the Million Veteran Program, VA’s largest research program on genes and health. VA also boasts a well established research infrastructure including clinical, epidemiological, imaging, and genetic expertise with well-established close partnerships with clinical operations. Further, compared with others in the US, Veterans enrolled in VA care experience a greater burden of multimorbidity and have higher rates of suicide and drug overdose.

DOE and its national labs have dramatically advanced artificial intelligence through world-class supercomputers, cutting-edge algorithms and secure computing infrastructure with robust software stacks such as through the Exascale Computing Program.^2^ With a leading scientific and technical workforce, DOE has delivered impactful solutions addressing other critical challenges including quantum computing, energy infrastructure, computing infrastructure and applied mathematics. DOE’s state-of-the-art capabilities for both open and secure research and development, coupled with a highly skilled workforce and a track record of strong partnerships across industry, academia, nongovernmental organizations and other federal departments and agencies promises mission-oriented innovation.

The first phase of MVP-CHAMPION established the foundation by securing interagency agreements, regulatory frameworks, high-performance secure computational environments that are certified to handle sensitive data, and proof-of-concept projects demonstrating feasibility and value.^3^ Technical lessons learned have been previously summarized.^3^ The group also rapidly pivoted to respond to acute information needs during the COVID pandemic.^4^ Here, we focus on Phase 2 of the collaboration, which strategically expanded the portfolio to eight new projects targeting high-burden conditions and unmet clinical needs (**Table 1 and Supplement**).These projects not only produced significant scientific findings—including discovery of novel genetic variants, development of predictive risk models, and creation of reusable AI and machine-learning (ML) frameworks—but also accelerated translation toward clinical application within and beyond the VA healthcare system.^5 6^

**Table 1.**
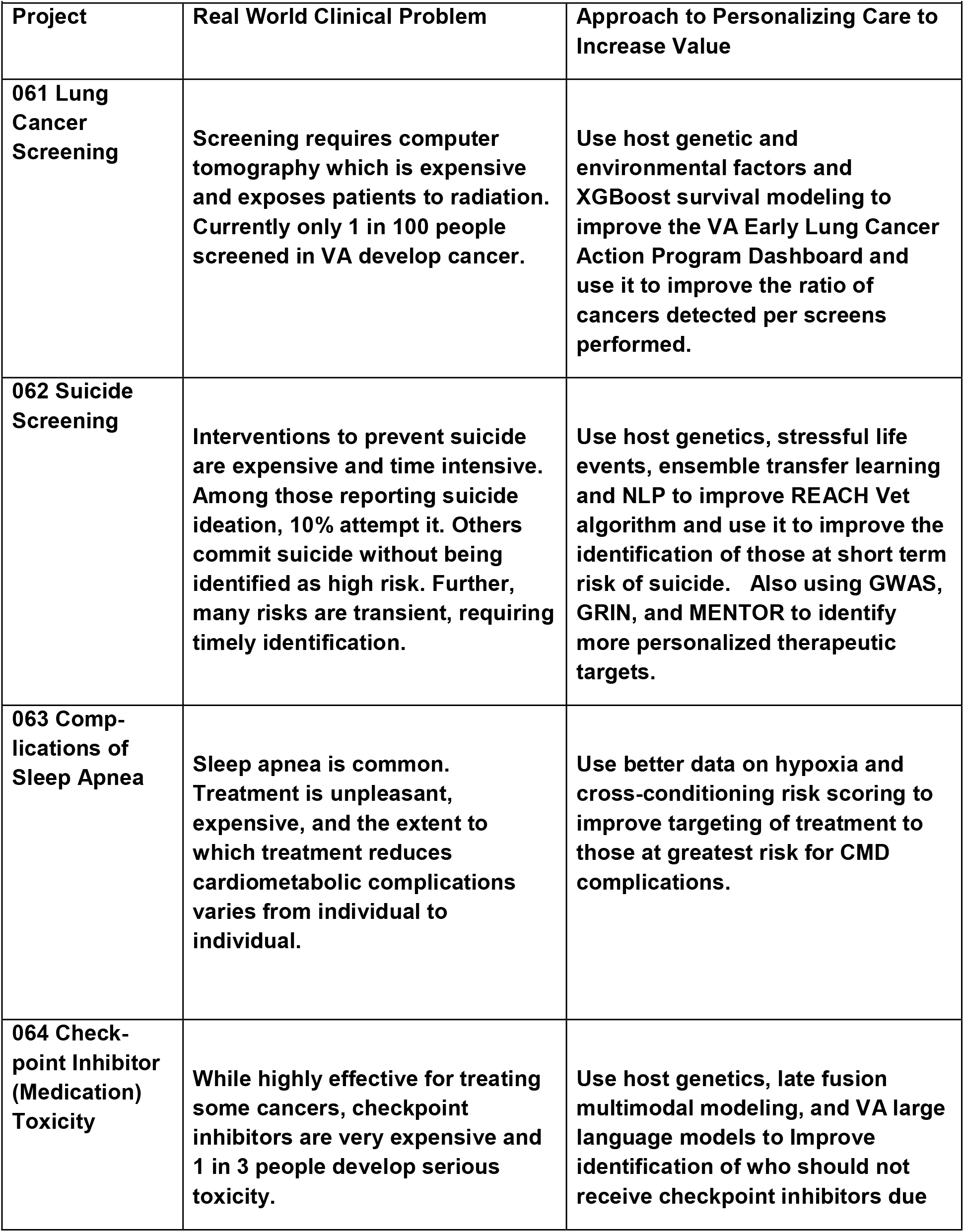
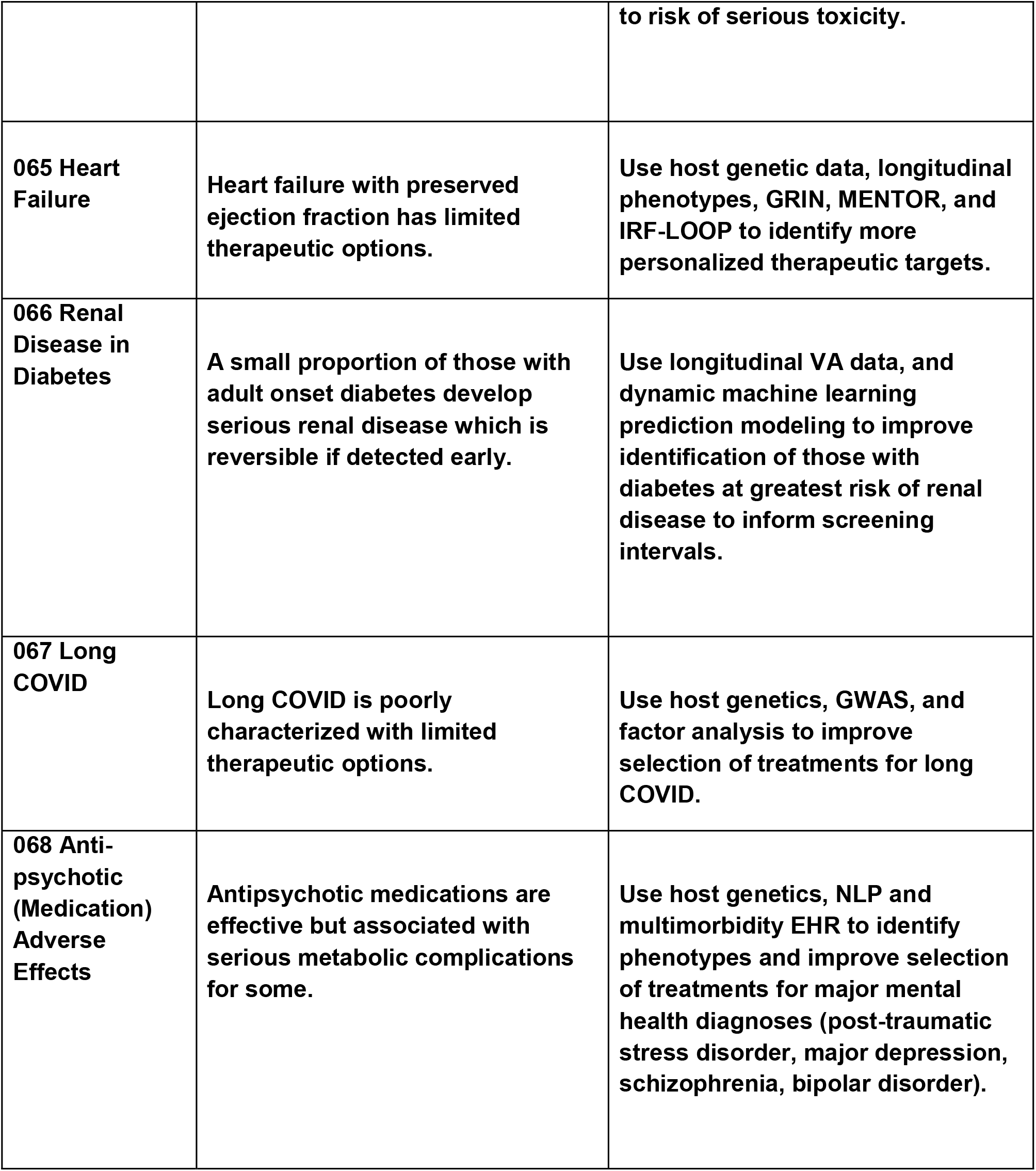
2nd Wave of MVP-CHAMPION Projects

## OBJECTIVE

This white paper summarizes the structure, activities, and impact of the Phase 2 MVP-CHAMPION projects. We highlight scientific discoveries, advances in predictive modeling and AI, operational efficiencies, and the broader vision for scaling precision health approaches across the VA—and beyond. By integrating discovery science with real-world clinical operations at national scale, MVP-CHAMPION offers a blueprint for how federal collaboration can lead to better science and to better health for millions of Americans.

## MATERIALS AND METHODS: VA-DOE A UNIQUE COLLABORATION

Several features distinguish the scientific approach of MVP-CHAMPION: A defining feature is its design around multimorbidity, recognizing that Veterans often experience multiple interacting conditions rather than isolated diseases. Unlike traditional single-disease studies, this initiative embraces the interconnected realities of patient health to produce models and interventions that reflect actual clinical complexity. It also acknowledges that Veterans’ health is influenced by an abundance of social and environmental determinants of health. Projects across the portfolio benefited from a shared computational infrastructure, including both cross-cutting tools, the gathering and curation of multimodal datasets, and a repository of carefully-defined predictor variables. These methods and data were reused and adapted across disease areas, reducing redundancy, promoting interoperability, improving validation, and accelerating time to impact.

To ensure translational relevance, clinical-operational integration was embedded from the outset. Research teams worked closely with VA clinicians and operational leaders to define outcomes, shape analytic approaches, and align research outputs with clinical workflows, paving the way for real-world application. Finally, a strong emphasis was placed on applicability across ancestry groups, particularly in genomics projects. Analytical strategies were designed to validate findings in populations across ancestral populations, ensuring that the benefits of precision health reach all Veterans equitably.

VA clinical expertise is central to the model, with operational leaders, practicing clinicians, and clinical researchers contributing a deep understanding of Veteran health needs and the realities of delivering care within the VA system. This expertise is coupled with VA data assets that include 30 years of longitudinal, largely paperless electronic health records (EHRs) from more than 25 million Veterans, linked to genomic and biospecimen data through the Million Veteran Program (MVP).

These resources are paired with the computational strengths of DOE, including access to AI-ready high-performance secure computing environments capable of analyzing petabyte-scale datasets, along with specialized expertise in AI/ML), and systems modeling.

Each project team is jointly led by VA and DOE investigators, ensuring that domain knowledge, computational innovation, and translational relevance are all embedded from the outset. This structure enables not only rapid discovery but also the practical alignment of research outputs with the clinical needs and realities of the VA healthcare system. It also enables iterative collaboration: clinical questions are informed by data science capabilities, computational innovations are guided by clinical needs, and outputs are tested with an eye toward operational feasibility and translation.

Moreover, the infrastructure enabled the development of a secure exascale AI capability (Frontier) that resulted in technologies that can contribute to scientific applications central to the DOE mission. The environment was deliberately designed to be disease-agnostic and enabling scale-out capabilities flexibly to simultaneously address a wide spectrum of health challenges, including suicide prevention and cancer care, agency priorities for VA and for DOE. Across Phase 2, the collaboration established a flexible and extensible ecosystem that supports:

- **Secure data access encompassing broad teams across the VA and DOE Labs**: Allowing real-world data analysis with stringent privacy and security standards.
- **Shared AI/ML Infrastructure**: Developed, adapted and reused novel methods GRIN^7^ (gene refinement), MENTOR^8^ (functional clustering), iRF-LOOP^9–11^ (predictive EHR modeling), VA-LLM (VA-trained large language model) and VISION (medical imaging ecosystem)^12,13^.
- **Rapid Cross-Project Transfer of Tools**: Innovations developed for one condition (e.g., suicide prediction models) were rapidly adapted for others (e.g., sleep apnea and diabetes progression modeling), maximizing efficiency.
- **Access to Supercomputers:** DOE’s leadership-class computing resources enabled analyses at a scale and speed not otherwise achievable in healthcare research^14^.
- **Domain specific/Biomedical AI workflows:** incorporating domain knowledge to guide AI model selection, data processing and decision making into pipelines that can be executed automatically (e.g. clinical note processing with LLMs).

We enabled transformative collaboration by fostering synergy across agencies, disciplines, phenotypes, data modalities, and spatio-temporal scales. Key elements of our approach included:

- **Establishing inter-agency agreements** that laid the foundation for coordinated action and shared objectives.
- **Building a large-scale, integrated team science model** that emphasized synergistic co-design and joint problem-solving.
- **Harmonizing data science and healthcare data** to unlock novel insights across domains.
- **Engaging researchers across multiple projects**, promoting the cross-fertilization of ideas, methods, and innovations.
- **Adopting a holistic framework** that addressed complex health challenges, including the impact of multimorbidity on outcomes.

This integrative approach lowered traditional barriers and catalyzed a new level of interdisciplinary innovation.

## RESULTS: PORTFOLIO OVERVIEW AND CROSS-CUTTING ADVANCES

The Phase 2 MVP-CHAMPION project portfolio collectively addresses critical areas of Veteran health—including suicide prevention, heart failure, lung cancer screening and treatment, diabetes complications, sleep apnea, post-acute COVID-19 sequelae, and antipsychotic-induced metabolic effects (**Table 1, Supplemental Materials**). They reflect a balance of early-stage discovery and translational readiness, demonstrating how foundational biological insights, real-world EHR data, and secure high-performance computing and supercomputing infrastructure can be integrated into a cohesive precision health pipeline. This overview illustrates not only the scientific depth of individual projects but also the strategic coherence of the overall portfolio in building a flexible, scalable framework for improving Veteran care.

While each project targets improving value in a distinct clinical domain, they collectively demonstrate the power of shared infrastructure, multimodal data integration, and a focus on solving real-world healthcare challenges through precision health. Common elements observed across projects include strong genetic drivers, the importance of health behaviors and substance use, multimorbidity, and the need to properly assemble the longitudinal and multimodal data sets with study designs appropriate to produce robust, actionable, and explainable conclusions. Insights gained in one project were shared across the collaboration.

Beyond the achievements of individual projects, Phase 2 of MVP-CHAMPION demonstrated the extraordinary power of integrated science when collaboration, shared infrastructure, and strategic reuse of methods are intentionally prioritized. Several major advances emerged across the entire portfolio, amplifying the scientific impact, operational efficiency, and translational readiness of the program as a whole.

Projects leveraged and iteratively improved a set of foundational computational tools:

- **VA Large Language Model (VA-LLM) enabled the extraction of clinical insights** from billions of clinical notes and they are being applied to the suicide prevention, lung cancer projects and antipsychotic adverse effects, as well as other VA projects not discussed here.
- **GRIN (Gene set Refinement through Interacting Networks) refined gene discovery** by integrating multiplex network connectivity. Already used across heart failure and suicide projects, GRIN are also be used in the long COVID and lung cancer projects to improve the biological relevance of genome-wide association study (GWAS)-derived gene sets.
- **MENTOR (Multiplex Embedding of Networks for Team-Based Omics Research) enabled identification of functional clusters of genes** from heterogeneous biological networks was applied to heart failure and suicide analyses.
- **iRF-LOOP (Iterative Random Forest Leave-One-Out Prediction) demonstrated the flexibility of explainable AI** frameworks across conditions. Originally developed for heart failure, iRF-LOOP has also been used to determine environmental and socio-demographic predictors of suicide attempts^11^ and could be used for other modeling projects modeling important health conditions.

Several projects accelerated progress by repurposing computational models and pipelines developed in related domains:

- **Accelerated Population Genomics Pipelines:** We engineered high-performance, GPU-accelerated workflows that seamlessly scale from cohorts of a few thousand individuals to over one million samples across thousands of phenotypes, enabling near-real-time discovery of genotype–phenotype associations, rapid hypothesis testing, and accelerated insights into complex trait architecture.
- **Precision Outreach Framework**: Originally developed for lung cancer screening, this framework could inform cardiometabolic disease outreach models (e.g., addressing statin undertreatment in screened Veterans) and has potential applications in suicide prevention and diabetes management.
- **NLP pipelines for risk factor extraction**^15^: Methods to extract social determinants of health (e.g., housing instability, food insecurity, social isolation, and troubles with the law) from unstructured text were first developed for the suicide project and subsequently adapted to extract concepts associated with obstructive sleep apnea and six of the mostly commonly associated comorbities and to extract life stressors associated with poor surgery outcomes. NLP methods using zero shot approaches were developed to extract social determinants of health and used in psychiatric adverse event prediction. These methods could also be adapted for other disease modeling such as diabetes.
- **Dynamic risk prediction models**: Joint modeling approaches built for diabetes complication prediction, antipsychotic adverse effect prediction can now be generalized to other progressive diseases such as heart failure and chronic lung disease.

A core principle across MVP-CHAMPION was the need to model multiple interacting conditions rather than treating diseases in isolation. Projects integrated multimorbidity explicitly by:

- **Embedding comorbidity variables** into predictive models for suicide, diabetes complications, lung cancer screening, and obstructive sleep apnea.
- **Using network models that captured genetic, clinical, and environmental interactions** across multiple organ systems.
- **Modeling influence of comorbid conditions** on risk, prognosis, and therapeutic needs in real-world populations.

Operational and translational synergies were achieved by working within an integrated VA-DOE environment:

- **Both high performance (HPC) and exascale computing were utilized**. While the bulk of the collaborative work occurred in a secure HPC environment, both genome-wide phenome-wide association analyses and natural language processing required exascale computing resources, with approximately 1,000 times the compute power of the HPC environment. Scaling the needed applications to this extent (PMC11142062) leveraged capabilities created through DOE’s exascale computing project (https://www.exascaleproject.org/). Without this resource, numerous smaller efforts would have produced less-integrated results.
- **Seamless and secure access to supercomputing resources was enabled**. The uniqueness of highly utilized leadership computing platforms necessitates novel encryption techniques to protect data. The collaboration implemented a secure infrastructure, Citadel, together with workflows designed to protect Veteran healthcare data while utilizing exascale computing. (https://researchcomputing.princeton.edu/systems/secure-research-infrastructure).
- **VA clinical and operations teams were engaged early**. Operational partners helped shape clinical relevance, outcome definition, and feasibility for real-world implementation from the inception of projects.
- **Cross-agency expertise was maximized**. Data scientists, genomicists, clinicians, and policy experts worked collaboratively, producing outputs that balanced scientific innovation with real-world applicability.

Taken together, these cross-cutting advances demonstrate that MVP-CHAMPION Phase 2 was not simply a collection of parallel research projects, but an integrated national effort to redefine what is possible when federal agencies coordinate data, computation, and mission around improving Veteran health.

## DISCUSSION: LESSONS LEARNED AND FUTURE DIRECTIONS

Phase 2 of MVP-CHAMPION confirmed that strategic alignment of data, computation, and multidisciplinary expertise can not only generate new scientific knowledge but also produce tools, models, and insights directly ready for clinical application. The lessons learned here, about integration, infrastructure, multimorbidity, equity, and translation, provide a blueprint for scaling this approach nationally and sustaining its impact over time. The success of Phase 2 of MVP-CHAMPION demonstrates that federal collaboration, when intentionally structured around real-world clinical needs, high-performance computation, and scalable data assets, can meaningfully accelerate both scientific discovery and health system transformation. Yet Phase 2 represents not an endpoint, but a foundation, a first proof that a new paradigm for precision health is achievable at national scale. Building on this momentum, we envision a future in which the VA-DOE collaboration continues to evolve into a flagship national model for personalized, data-driven, preventive, and efficient healthcare. Efforts towards these goals are actively ongoing.

1. **Federal integration and collaboration is essential**. The most successful projects embedded VA clinicians and researchers, DOE data scientists, DOE computational biologists, and VA operational leaders into a single, iterative workflow. Close integration ensured that clinical questions were framed in a computationally tractable way, analytic methods were chosen with real-world deployment in mind, and outputs were immediately interpretable by clinicians. Early and ongoing collaboration across disciplines accelerated both scientific innovation and translational readiness. The VA-DOE collaboration provides a scalable blueprint for larger national efforts in the future. By expanding partnerships with other federal agencies, healthcare systems, and industry collaborators, MVP-CHAMPION can help accelerate discovery, validation, and deployment of precision health solutions not only for Veterans, but for the American public at large.
2. **Building a flexible, reusable secure infrastructure and pipeline for implementation saves time and costs**. Investments in shared computational assets, such as the VA Large Language Model (VA-LLM), the GRIN, VISION, and MENTOR network analysis tools, and common EHR pipelines, dramatically reduced project startup time, eliminated duplication of effort, and allowed knowledge and methods to flow across disease areas. Projects that could immediately leverage these assets progressed more rapidly from concept to discovery to translation. Many of the predictive models, AI tools, and computational pipelines developed in Phase 2 are now poised for broader operational deployment. In the future, expanding precision outreach frameworks, dynamic risk prediction dashboards, and multimorbidity-aware clinical decision support tools across the VA system could deliver immediate improvements in Veteran health, particularly for high-burden conditions such as heart failure, lung cancer, and diabetes.
3. **Modeling Multimorbidity Is Critical for Real-World Relevance** Projects that explicitly incorporated multimorbidity into their models consistently produced findings that were more predictive, clinically meaningful, and operationally actionable. Veterans rarely present with isolated disease states; efforts that acknowledged the complex, interacting nature of multiple conditions were better aligned with actual clinical realities and provided more accurate risk stratification and intervention targets. Phase 2 made substantial advances in integrating structured EHR data, unstructured clinical notes, genomics, and environmental data. The next frontier is seamless fusion across even more modalities—imaging, wearable device data, social determinants of health—to model the full complexity of Veteran health. Moreover, expanding multimorbidity modeling from a secondary consideration to a primary analytic focus will better reflect real patient trajectories and therapeutic needs. The emergence of digital twin technologies, computational representations of individual patients that evolve with longitudinal multimodal data, offers a powerful opportunity to simulate disease progression and assess personalized interventions. These virtual patient models can help test therapeutic strategies in silico, reducing risk and accelerating translation into clinical practice.
4. **Exascale Computing Unlocks Healthcare Scale and Complexity** Analyses of the scale required—encompassing millions of Veterans, multimodal data, billions of clinical notes, and multiscale biological data—would not have been feasible without access to DOE’s leadership-class computing infrastructure. High-throughput modeling, large-scale machine learning, and integration of structured and unstructured data all depended on DOE’s computational environment and expertise. Future expansion will continue to rely on scalable HPC and AI ecosystems. As healthcare data continues to accumulate, static risk models rapidly become outdated. Future MVP-CHAMPION efforts will aim to focus on creating real-time, continuously learning systems, adaptive models that update dynamically with new EHR entries, genomic discoveries, environmental exposures, and treatment innovations. This will move VA closer to a true learning health system where prediction and intervention evolve alongside clinical practice.
5. **Early Attention to Operational Translation Increases Impact** Projects that engaged operational partners from the outset, such as the Suicide Risk prediction effort, were better positioned to move from discovery to deployment. Benefits of early engagement were numerous, including assessments of fruitful directions for improvement, understanding of existing processes, and detailed discussions of appropriate validation strategies and problem formulations which produce actionable information and scalable implementation.
6. **The VA Continues to be a Valuable Source of Multi-Ancestry Genomic Analyses** As found previously within MVP, Genetic discoveries validated across ancestry groups were more robust and generalizable. Additionally, 12% of identified genomic risk loci in the MVP data were only significant in people of non-European ancestry (Verma, ref 14 below). Future phases must continue to prioritize within and cross-ancestry analyses, validation across demographic groups, and ensuring that precision health benefits extend to all Veterans, regardless of background or service history. Investment in equitable data collection, inclusive model development, and focused deployment in underserved populations will be critical.
7. **Administrative and Regulatory Alignment Enables Speed** The Interagency Agreement between VA and DOE, along with clear regulatory frameworks for data sharing and security, was foundational. All projects leveraged pre-approved environments, such as the VA KDI environment at the Oak Ridge National Laboratory S and DOE’s exascale computing facilities with appropriate authority to operate (ATO), were able to move faster without compromising privacy or compliance. The healthcare challenges of the next decade will require even greater computational innovation. Continued collaboration with DOE will ensure that VA researchers and clinicians can access next-generation AI methods (e.g., federated learning, explainable multimodal transformers), petascale-to-exascale computing, and cutting-edge simulation tools to push the boundaries of predictive, preventive, and personalized medicine.

## CONCLUSION

The MVP-CHAMPION collaboration between the VA and DOE is a broad and systematic effort to apply HPC and novel AI methodologies to develop precision health at national scale. The first example of success in this effort is the incorporation of new risk factors for suicide and deployment of the REACH-VET 2.0 tool across the VA healthcare system. Our experiences, described here, demonstrate the value of integrating clinical need, data science, and high-performance computing. Phase 2 delivered new discoveries, translational tools, and operational efficiencies that directly advance Veteran care. By continuing to expand, adapt, and innovate, MVP-CHAMPION is poised to serve as a national model for how federal science can drive real-world healthcare transformation, improving lives while setting new standards for impact-driven, collaborative precision medicine. A key insight emerging from this phase of work is the importance of supporting the full spectrum of technology readiness, from foundational biological discovery to late-stage clinical deployment. Many projects began with complex, high-resolution clinical data and used it not only to power near-term predictive models but to reveal core mechanistic signals, such as previously unrecognized genetic variants, pathway-level relationships, or new physiological intermediaries, that are essential to next-generation therapeutics and diagnostics. This capacity to move fluidly between data science, basic biology, and clinical insight was made possible by the integrated structure of the collaboration and should be preserved and strengthened in future phases.

Equally important is the recognition that the clinical needs driving these projects are often more complex than they appear. For example, in suicide prediction, the highest-risk patients identified by models may not always be those who are most reachable or responsive to intervention. Similar patterns of intervention complexity and unintended trade-offs emerge in other domains, from cancer screening to cardiometabolic risk management. These examples underscore that precision health is not just about better risk prediction, it is also about understanding when, how, and for whom those predictions can lead to meaningful clinical benefit. Taken together, these experiences emphasize the need for ongoing, iterative interaction across the translational continuum, not only to move discoveries forward but to feed practice-based insights and edge cases back into the design of data pipelines, analytic strategies, and foundational scientific questions. This loop of co-design and co-evolution across basic science, computational modeling, and clinical operations is a defining strength of MVP-CHAMPION and should remain a core tenet of its future vision.

## Data Availability

All data produced in the present study are available upon reasonable request to the authors

## Acknowledgements

This manuscript has been authored in part by UT-Battelle, LLC, under contract DE-AC05-00OR22725 with the US Department of Energy (DOE). The US government retains and the publisher, by accepting the article for publication, acknowledges that the US government retains a nonexclusive, paid-up, irrevocable, worldwide license to publish or reproduce the published form of this manuscript, or allow others to do so, for US government purposes. DOE will provide public access to these results of federally sponsored research in accordance with the DOE Public Access Plan (http://energy.gov/downloads/doe-public-access-plan).

## SUPPLEMENT: SECOND WAVE PROJECTS

**MVP-CHAMPION 061: Lung Cancer Screening**

**VA Lead: Samuel Aguayo**

**DOE Lead: Ioana Danciu**

### Rationale

Approximately 7,000 U.S. Veterans are newly diagnosed with lung cancer each year. Because most are at a late stage and likely to die within 1-2 years, there is an increasing call for widespread screening to detect lung cancer at an earlier stage, when it is still curable. Over 200,000 Veterans are actively participating in lung cancer screening, but another 800,000 have yet to be enrolled and only 1% of those screened are diagnosed. This project aims to enable more efficient and early lung cancer screening, while screening fewer Veterans overall. Of note, the average spending by the VA on the treatment of late-stage lung cancer is estimated at more than $300,000 per patient. Surgical cure of early-stage lung cancer is estimated at less than $40,000 per patient.

### Aim

This project aims to refine the VA Early Lung Cancer Action Program (ELCAP) Dashboard, a clinical informatics tool that interrogates the electronic health record (EHR) to list Veterans at high risk, to “pull” high-risk Veterans into lung cancer screening using a process named “precision outreach”. As we identify new risk factors from diverse data sources and modalities, each revealing unique aspects of risk, the risk-based multimodal model can be integrated with the ELCAP Dashboard for validation in real time across VA facilities, and for real life prospective use in clinical operations.

### Data

We used the entire VA population with visits in 2016 (4,893,984 patients). Our outcome was lung cancer diagnosis in the interval 1/1/2017-1/1/2021 (23,660 patients). Our predictors included demographics, history of pulmonary and cardiac disease, environmental exposures to air pollution, and medications. We also performed genome-wide association studies (GWAS) on 3,733 lung cancer cases and 41,106 cancer-free controls of African, 17,995 cases and 112,343 controls of European, and 768 cases and 21,119 controls of Hispanic ancestry with MVP genotyping.

### Analytic Methods

We used a machine-learning survival analysis approach using xgboost to predict time to lung cancer diagnosis. This required the high-performance computing infrastructure at Oak Ridge National Laboratory. We split our dataset into stratified train, validation, and test sets with similar outcome prevalence. Our metric was Harrell’s c-statistic and we report it on the test dataset, set aside before training. For increasing the translational impact, we also analyzed the predictor importance of the model features.

### Results

The c-statistic for this model was 0.81 on the test set, and the most significant features were confirmed to be patient age, tobacco use, and chronic obstructive pulmonary disease. Environmental air pollution data was the next stronger predictor of risk for lung cancer, but such data is not readily available in the EHR, and it was only possible to learn about it because of the computing infrastructure available through this collaboration with DoE. The GWAS identified potentially new risk common variants associated with lung cancer.

### Next Steps/Application/Impact

We are working on independent validation and will be adding genetic risk factors to the ELCAP Dashboard, leveraging the existing VA Pharmacogenomics EHR infrastructure.Our work focuses on identifying predictors of lung cancer risk and spans from early translational phase to direct translation into the clinic and health care decision making once the new discoveries are added to the ELCAP Dashboard. There are current collaborations with MDClone, another VA contractor with authority to operate in VA ARCHES, for rapid translation of VA ELCAP Dashboard and insights from our work into additional tools for both additional research and for improving day to day clinical operations. Several publications forthcoming from these collaborations.

### Efficiencies Gained

Refining the VA ELCAP is only possible through group collaboration. Its development and current application are possible by shared expertise and infrastructure across multiple projects in the collaboration. In addition to lung cancer screening, many complex health care challenges can benefit from precision outreach using tools like the VA ELCAP, and the adaptation of our tools and procedures becomes a frequent topic of communications when we present to different audiences.This work would not have been possible without the high-performance computing expertise and infrastructure from the Oak Ridge National Lab team, and the genetics computational expertise of the MVP investigators.

### Role of Multimorbidity

Accounting for multimorbidity is essential in developing accurate models for improving survival from lung cancer screening, and likely a reason why no trials have significantly decreased all-cause mortality despite the dramatic results in lung cancer specific mortality. Most lung cancer patients present with comorbidities such as atherosclerotic cardiovascular disease and pulmonary disease, that can significantly affect outcomes. For instance, although atherosclerotic cardiovascular disease is not a strong predictor of lung cancer risk, with our ELCAP Dashboard we found that most Veterans in lung cancer screening have increased atherosclerotic cardiovascular disease risk and have atherosclerotic cardiovascular disease events while participating in lung cancer screening. Furthermore, we have also found that about 30% of Veterans currently participating in lung cancer screening have increased low density lipid levels that warrant cholesterol reducing intervention but do not have an active prescription for a statin in the EHR. Using “precision outreach” we are addressing the undertreatment of atherosclerotic cardiovascular disease risk during lung cancer screening at Phoenix and working with the VA National Center for Patient Safety to take action across VA.

**MVP-CHAMPION 062: Suicide**

**VA Lead: Jean Beckham**

**DOE Lead: Silvia Crivelli**

### Rationale

The rate of suicide among U.S. Veterans increased by 65% from 2001 to 2022^1^. As a result, the sex- and age-adjusted suicide mortality rate for Veterans is now 58% higher than that of non-Veterans^2^. In 2022 alone, 6,407 Veterans died by suicide, making it the 12th leading cause of death among all Veterans and the 2nd leading cause of death for Veterans under the age of 45^3^. A recent analysis estimated the annual economic costs associated with suicide and non-fatal self-harm to the United States to be more than $500 billion dollars per year^2^.

### Aim

To address the pressing concern of Veteran suicide, the VA-DoE partnership supported multiple studies aimed at identifying risk factors for suicide among Veterans^1,4–14^. Among other accomplishments, these studies have resulted in the identification of novel genetic risk loci for suicide; the identification and extraction of stressful life events from the unstructured data; the identification of social and environmental determinants of health; new computational methods for analyzing genomic data; and increased understanding of the biology of suicide^4–17^.

### Data

The ensemble transfer learning and deep neural network models used structured EHR data that included all 24,558,158 distinct patients for whom a date of birth was available on or after January 1, 1901. Natural Language Processing (NLP) and large language models (LLMs) used unstructured EHR data, which include 4,200,093,996 ReportTexts generated from 4,354,267,392 care encounters for 14,062,540 patients (national PatientICN). Geospatial data were gathered from multiple publicly available sources, including Agency for Healthcare Research and Quality, American Community Survey, County Health Rankings and Behavioral Risk Factor Surveillance System, Met Office Hadley Centre observations^18^, and National Oceanic and Atmospheric Administration.

### Analytic Methods

These projects have identified numerous non-genomic risk factors for suicide and suicidal behavior through the application of a wide range of machine learning approaches, including iterative random forest, NLP, ensemble transfer learning, and deep sequential neural network models^1,11–13^.

### Results

NLP was used to identify stressful life events (e.g., homelessness) from clinical notes associated with risk for suicide that would be underestimated if only structured data was to identify risk factors^13,19^ for suicide. To that end, we developed a NLP pipeline that performs named-entity recognition over all VA clinical text and discriminates false-positive annotations to reduce clinical burden. The pipeline runs with high performance computing in the backend to support concept extraction across all the VA clinical text^15^. Ensemble transfer learning models were used to successfully predict death by suicide, suicide attempt, and overdose diagnoses in a prospective cohort of 4.2 million Veterans^11^. Additionally, a preliminary study of deep sequential neural networks found that these methods resulted in substantially higher accuracy for the prediction of future suicide attempts than traditional approaches^12^. For example, more than 99% of the Veterans ranked in the upper 0.1% risk tier by our top deep sequential neural network model attempted suicide during the following year in the test set, translating to an adjusted positive predictive value of 0.54^12^.

### Next Steps/Application/Impact

We are currently focused on expanding this work using deep sequential neural networks to predict suicide deaths in the total VA cohort. It should also be noted that there are now multiple predictors that have identified by the suicide exemplar team (e.g., food insecurity, changes in marital status, NLP-based prediction of homelessness) that have been implemented by our partners in VA clinical operations in order to improve VA’s ability to prevent suicide among high-risk Veterans.

### Efficiency Gained

Many of the methods that were developed as part of our initial suicide project to overcome challenges inherent to the study of suicide (e.g., extremely low base rates) have been critical to the success of other projects.

### Multimorbidity

To maximize statistical power, models were initially trained to predict combined outcome of death by suicide, suicide attempt, and overdose. Most patients found in the top strata of structured-based models have psychiatric diagnoses. According to Coon et al. 80%-90% of those patients will not die by suicide^20^. Preliminary results from the VA-LLM show another group of patients with absence of non-fatal suicidal behavior among those at the top 0.1% stratum. They show conditions such as chronic pain, spinal cord injuries, and cancer.

**MVP-CHAMPION 063: Sleep Apnea VA**

**Lead: Ellis Boudreau**

**DOE Lead: Patrick Finley**

### Rationale

Untreated Obstructive Sleep Apnea (OSA) is highly prevalent in Veterans (24%) and is associated with cardiometabolic disease. While there is some correlation between OSA severity and susceptibility to cardiometabolic disease, high inter-individual variability makes it difficult to identify those individuals at highest risk for disease and in need of targeted therapy^21^. Furthermore, adherence with the primary therapy, Continuous Positive Airway Pressure (CPAP) is low and the costs of therapy and interventions for improving adherence are resource intensive^22^. Because multiple pathophysiologic pathways contribute to the development of OSA and related co-morbidities, efforts to-date have been hampered by the large number and breadth of data features that need to be analyzed to develop tools that not only predict the risk of developing cardiometabolic disease but also the time course of cardiometabolic disease development. However, many of these obstacles can be overcome using predictive AI-based models, high performance computing, and the large amount of data in the EHR.

### Aim

We set out to develop a predictive analytic tool for identifying those individuals at highest risk for developing cardiometabolic disease from OSA and to determine the temporal pattern of occurrence.

### Data

We analyzed EHR data which includes structured and unstructured data for over 23 million Veterans, spanning 25 years. At the time of analysis, records were available between 01/01/2000 and 12/31/2022. Using International Classification of Diseases (ICD) and Current Procedural Terminology (CPT) codes, we identified 2.6 million Veterans with OSA and we selected a subgroup of 1.1 million Veterans who had either a home or in-lab sleep test within the VA. Most Veterans in the OSA positive group developed cardiometabolic disease either prior to or after their sleep test demonstrating the high prevalence of cardiometabolic disease in our population of interest. Veterans in this cohort were required to have data in their medical record for at least two years prior to their sleep test to ensure that cardiometabolic disease pre- and post-testing were captured. For example, if a Veteran had a sleep test in January 2002, then data on cardiometabolic disease development would be available for 2 years prior to their sleep test and for 20 years after it.

### Analytic Methods

We developed a pipeline for the analysis of multiple cardiometabolic diseases with overlapping risk factors for use in studies using large clinical databases, and NLP and machine learning (ML) approaches. The pipeline was also used to extract stressful life events for the suicide exemplar. This approach demonstrates the reusability of computational infrastructure across a diverse set of disease models. A predictive logistic regression model incorporating data from both the Veterans with OSA and without OSA was used to determine the likelihood of developing a new cardiometabolic disease after sleep testing (T0).

### Results

The results from this model demonstrate that the odds for developing a new cardiometabolic disease after T0 were 2.13 with an area under the receiver operating characteristic of 0.74, which is consistent with the literature showing that OSA is associated with an increased risk for cardiometabolic disease. Currently, we are developing deep learning models that predict the future occurrence and timing of a possible cardiometabolic disease event in Veterans with OSA.

### Next Steps/App/Impact

We are extending this foundational work into the role of OSA in modifying the development of cardiometabolic disease to the study of how military toxic exposures and OSA may co-modulate the development of cardiometabolic disease. We are also creating a beta version of a program to predict cardiometabolic disease risk for use in sleep clinics; next steps - refinement of the program, scale it for use on VA computer systems; and testing. Our tool could be adapted for use in active duty military to help assess risk of apnea complications during deployment, and is an example of how predictive analytics could be used to assess troop medical readiness. Further, the amount of hypoxia seen during apneic episodes appears to be a major driver for OSA associated cardiometabolic disease(hypoxia phenotype). We are using models developed for this project to inform a collaboration with the military to evaluate hypoxia unawareness in aviators, which remains a significant operational issue and contributes to aviation mishaps.

### Efficiencies Gained

For our project, both the NLP pipelines and ML work were informed by approaches developed to enhance the prediction of suicide using the tool suicide exemplar. The infrastructure created for the suicide exemplar will also be utilized for the deployment of our tool. Additionally, we used both validated (obstructive sleep apnea) and machine generated phenotypes (heart failure, hypertension, stroke, atrial fibrillation, coronary artery disease, and diabetes mellitus type 2) from the VA Centralized Interactive Phenomics Resource as a starting point for identifying cardiometabolic disease associated with OSA.

### Role of Multimorbidity

This project looked at six of the most common comorbidities associated with sleep apnea, clinically it is not all that useful to just look at one or two co-morbidities as you really need to be able to assess risk across a common set of co-morbidities, so this aspect was absolutely critical to clinical utility of our project.

**MVP-CHAMPION 064: Lung Cancer Treatment**

**VA Lead: Michael Green and Alex Bryant**

**DOE Lead: Shinjae Yoo**

### Rationale

Approximately 1,000 to 2,000 Veterans receive immunotherapy with checkpoint inhibitors to treat non-small cell lung cancer (NSCLC) annually and one in three will experience significant immune-related toxicity from these treatments. Use of checkpoint inhibitors to treat various forms of cancer is rapidly expanding with recent annual spending by VA on immunotherapy drugs is approximately $540 million (60-70% of these costs are to treat NSCLC)^23^ Unfortunately, these toxic events are extremely unpredictable and are often highly morbid.

### Aim

Our work focuses on identifying basic mechanisms and predictors of immune checkpoint inhibitor (ICI) toxicity and efficacy.

### Data

Approximately 6,000 to 7,000 U.S. Veterans are newly diagnosed with non-small cell lung cancer (NSCLC) each year. While NSCLC accounts for approximately half of our study population, our research cohort includes a broader, pan-cancer population of Veterans with stage III–IV malignancies receiving immunotherapy. This expanded scope increases the potential impact of our work beyond NSCLC alone. The broader applicability of our predictive models suggests that the relevance of our findings will extend to a significantly larger population of Veterans undergoing immunotherapy across multiple cancer types.

### Analytic Methods

We used logistic regression and then applied late fusion techniques to integrate structured features and VA-Large Language Models (VA-LLM) activations. The VA-LLM consists of 1.62B parameters pretrained on 1.4T tokens of VA clinical text. We performed genome-wide association studies to identify genetic determinants of survival and immune-related toxicities. We also explored the influence of polygenic risk scores for autoimmune comorbidities on immunotherapy toxicity development.

### Results

Performance metrics resulting from applying logistic regression and the VA-LLM to the 180 days survival prediction (Table below). We also include the results using two open-source LLMs (Mistral-large and Llama 3.3). The VA-LLM was the most accurate predictor of our clinical outcomes despite being considerably smaller.we performed GWAS for ICI efficacy (survival) and toxicity, identifying potential novel genetic determinants (manuscript forthcoming). We also evaluated polygenic risk scores(PRS) and found that thyroid toxicity could be robustly predicted with a hypothyroidism PRS. Finally, we discovered a novel association between race and toxicity risk^24^ leading to multiple ongoing research projects to understand the mechanisms.

**Table.** Late Fusion Results. Mean Metrics Over K-Fold Cross Validation

**Table. Late Fusion Results. Mean Metrics Over K-Fold Cross Validation**
| Model | AUROC | Accuracy | F1 | Precision | Recall |
| --- | --- | --- | --- | --- | --- |
| Clinical Logistic Regression | 0.774 | 0.715 | 0.586 | 0.504 | 0.702 |
| VA-LLM | 0.768 | 0.715 | 0.586 | 0.504 | 0.702 |
| VA-LLM + Clinical Vars | **0.792** | **0.743** | **0.607** | **0.542** | 0.690 |
| Llama 3.3 70B | N/A | 0.646 | 0.574 | 0.443 | **0.814** |
| Mistral-Large 123B | N/A | 0.685 | 0.587 | 0.476 | 0.765 |

This table shows performance metrics resulting from applying late fusion techniques to integrate structured features and VA-LLM activations. The VA-LLM uses the last 30-day of deduplicated clinical text to predict outcomes. Logistic regression was used to build ensemble prediction between modalities.

In addition to training the VA-LLM from scratch on billions of clinical notes, we have developed a robust evaluation pipeline to benchmark the performance of open-source LLMs, such as LLaMA 3 and Mistral for the same clinical prediction tasks. This pipeline incorporates advanced prompt engineering strategies to enhance performance and minimize hallucinations. It is adaptable across a range of clinical applications, making it a foundational tool that supports other projects within the group.Finally, we identified several genomic loci associated with prolonged survival after immunotherapy treatment, and several loci possibly associated with immune-related adverse events (manuscript under preparation). We found that a polygenic risk score for hypothyroidism robustly predicted thyroid toxicity from immunotherapy. Finally, we discovered a novel effect of self-reported race on immunotherapy toxicity^24^.

### Next steps/Applications/Impact

We will next validate our predictive models in prospective settings and integrate them into clinical decision support tools.We are performing additional studies to confirm the influence of race on toxicity risk and planning additional work to better understand the mechanism.

### Efficiencies

This work would not have been possible without the high-performance computing expertise from the DoE Lawrence Berkeley Laboratory team, which enabled the training of the VA-LLM. Additionally, the multimodal machine learning expertise from the Lawrence Berkeley team was critical in integrating LLM outputs with high-dimensional clinical data to build effective predictive models.The VA-LLM, a large language model developed through group collaboration, was the most accurate predictor of our clinical outcomes. Its development and application were made possible by shared expertise and infrastructure across multiple projects in the collaboration.In turn, our VA-LLM, trained on billions of clinical notes and trillions of tokens, is a foundational model with applications to other areas in healthcare.

### Multimorbidity

Accounting for multimorbidity was essential in developing accurate models for predicting overall survival after immunotherapy. Most lung cancer patients present with comorbidities such as cardiovascular disease and chronic obstructive pulmonary disease, which significantly influence outcomes.

**MVP-CHAMPION 065: Heart Failure**

**VA Lead: Jacob Joseph**

**DOE Lead: Daniel Jacobson**

### Rationale

Heart failure, a clinical syndrome arising from diverse etiologies, is a major global health challenge that affects over six million people in the United States^25^. Among U.S. Veterans, the burden is even more severe: approximately 5% of Veterans have heart failure—more than double the 2.4% prevalence in the general population—and the condition carries an annual mortality rate of 14.5%^26^. The financial impact is substantial, with U.S. annual heart failure care costs projected to exceed $70 billion by 2030. Within the VA, the average annual expenditure attributable to heart failure has been estimated at $8.3 billion^27^.

### Aim

To apply an integrated, mechanistic approach to understanding the molecular and clinical heterogeneity of heart failure, with a particular focus on the difference between heart failure with reduced and preserved ejection fraction (HFrEF and HFpEF), the latter being a form of the disease that remains refractory to most existing therapies.

### Data

The analysis draws on a broad spectrum of data. EHR data, and echocardiography was used to define the baseline HFrEF and HFpEF phenotypes. EHR data and ML models (iRF-LOOP) were used to define the HFpEF sub-phenotypes. GWAS summary statistics for unspecified HF, HFrEF and HFpEF phenotypes supplied the initial list of SNPs. Three complementary SNP-to-gene strategies—nearest-gene proximity, Multi-marker Analysis of GenoMic Annotation (MAGMA) linkage-aware mapping, and High-throughput Chromosome Conformation Capture coupled-MAGMA (H-MAGMA) chromatin-contact mapping that leverages High-throughput Chromosome Conformation Capture (Hi-C) from cardiovascular, pulmonary and hepatic tissues—translated those variants to candidate genes. To evaluate how those genes interact biologically, we assembled a multilayer gene–gene multiplex. Its evidence layers included HumanNet v3 co-expression, protein–protein interaction, molecular pathway co-membership, genetic interaction, gene-neighbourhood and phylogenetic co-occurrence edges, plus a co-citation layer. Two additional regulatory layers captured transcription-factor binding derived from Encyclopedia of DNA Elements (ENCODE) and from Human Umbilical Vein Endothelial Cells (HUVEC) chromatin immunoprecipitation studies, while seven tissue-specific predictive-expression networks were built from bulk and single cell RNA-seq by the iRF-LOOP explainable-artificial intelligence (AI) algorithm for aorta, atrial appendage, left ventricle, coronary artery, whole blood/lymphocytes, lung and liver. Hi-C contact maps therefore contribute 3-D genome structure, Genotype-Tissue Expression Project (GTEx) provides quantitative transcriptomes, and ENCODE supplies transcriptional regulation. Finally, DrugBank annotations supply approved and experimental drug–target relationships that anchor the downstream therapeutic network analysis.

### Analytic Method

Our work has spanned the research continuum from gene discovery to functional interpretation and clinical stratification. Using GWAS from the Million Veteran Program (MVP), we developed GRIN (Gene set Refinement through Interacting Networks)^9^ and MENTOR (Multiplex Embedding of Networks for Team-Based Omics Research)^28^ to refine SNP-to-gene associations and functionally cluster genes using multiplex biological networks composed of protein interactions, regulatory elements, and predictive expression layers built via explainable-AI. In parallel, we applied iterative random forests leave one out prediction (iRF-LOOP)^29,30^ (a method originally developed for other DoE projects) and network-based clustering to electronic health record data from over 55,000 VA patients with HFpEF. This yielded reproducible sub-phenotypes defined by distinct comorbidity trajectories, organ system involvement, and treatment histories, directly addressing the clinical heterogeneity that has limited therapeutic progress in HFpEF.

### Results

By integrating HF-associated gene clusters with drug-target networks, we identified both approved and investigational therapies with potential for repurposing in heart failure. Targets include genes in pathways involving inflammation, fibrosis, cytoskeletal remodeling, and myocardial energetics.

### Next Steps/Applications/Impact

These findings have already prompted discussion with industry partners around preclinical validation and repurposing strategies.

### Efficiences

These methods—developed in coordination with other DoE projects, in particular the suicide project—were adapted and extended to accommodate the specific needs of heart failure genetics, saving development time and ensuring interoperability across teams. This collaborative structure not only allowed us to scale analyses efficiently but also ensured that methods and findings were immediately useful to others in the consortium, including those studying polygenic traits and multimorbidity in different disease domains.

### Multimorbidity

The ability to incorporate multimorbidity and transfer learning across conditions—enabled by training predictive expression networks in disease-relevant tissues— was essential in identifying biologically coherent signals across datasets.

**MVP-CHAMPION 066: Renal Disease in Diabetes**

**VA Lead: Peter Reaven**

**DOE Lead: Benjamin McMahon**

### Rationale

Diabetes affects nearly 25% of VA’s patient population and is a major cost-driver for the VA^31^. The total estimated cost of diagnosed diabetes in the United States in 2022 is $412.9 billion, including $306.6 billion in direct medical costs and $106.3 billion in indirect costs attributable to diabetes^32^. Diabetes has systemic effects and the types of complications that develop from diabetes are diverse and involve almost every organ and vascular system in the body. Adding to this complexity, many of these complications have additive or possibly synergistic effects on other outcomes. For example, although diabetes is the leading cause of end-stage renal disease, development of renal disease also increases risk for heart disease^33–35^. Importantly, many diabetes medications now available appear to slow the development and/or progression of renal disease in diabetes. Thus, improving our identification of persons with diabetes that are most likely to develop renal disease and need additional monitoring or therapy is critical to reducing morbidity with diabetes and concomitant financial cost.

### Aim

Our project seeks to integrate state-of-art ML/AI techniques with our newly developed scalable joint modeling methods to study individual health trajectories to improve disease prediction^36^.

### Data

This approach allowed us to combine diverse data types in EHR records (noted above) to dynamically predict the onset of end stage renal disease (ESRD) in patients after they were diagnosed with diabetes.

### Analytic Methods

We built a flexible modeling system that helped us develop and validate an ESRD risk prediction model for newly diagnosed diabetes patients, using data from two major EHR systems.

### Results

Our results demonstrated very good performance in the VA cohort with Area Under the Curve > 93% and good calibration (manuscript submitted). The dynamic (vs static) prediction model further improves prediction model’s calibration and is superior to the current state of the art ESRD risk scores. Moreover, we were able to demonstrate that variables describing the same ML-selected health attributes that predicted ESRD in the VA could be used effectively in an entirely separate set of health records collected from different patients (All of US cohort). Thus, the method was transferable to and validated in a conglomerate of different health systems. More work, sometimes described as federated learning, is required to systematize the process applying VA-developed models to alternative sources of data.

### Next Steps/Applications/Impact

As our real-world healthcare data reflect records of actual patients, this work has direct translational implications. With further refinement and validation, the prediction model could be deployed as a dashboard that extracts data and automatically updates risk prediction as new health data accumulate over time. These ML/AI approaches also provide robust estimations of the relative importance of risk factors and their changes to help guide intervention that may reduce future risk. As one example of the potential for this approach to encourage scientific collaboration and translational progress, Bayer Pharmaceuticals (Digital Health Division) has expressed interest in further defining and refining AI models to predict renal disease. The shared goal of these efforts is to identify patients within other health care institutions that are at higher risk for the entire spectrum of renal disease development.

### Efficiencies

The combined access to the VA’s extensive EHR data, essential for training our dynamic prediction model, and the DoE computing resources and AI/ML tools, which would have cost tens of thousands of dollars otherwise, made this project feasible. This support enabled us to develop a robust foundational prediction model. Additionally, the trained model facilitates transfer learning for future projects, allowing refinements without starting from scratch, therefore saving substantial computing costs. Further, building on prior expert work by DoE scientists provided an existing, high-performance computing optimized data processing pipeline. We then streamlined essential steps such as data pre-processing, quality control, and cohort definition, thereby significantly cutting project time and reducing potential errors. This strategic partnership ensured consistency in using real-world data and maximized efficiency by leveraging cutting-edge computing resources that would have been prohibitively expensive otherwise. Moreover, we developed a generalizable modeling framework, open-sourced on GitHub with comprehensive documentation, that not only can be used for the ESRD prediction model but also serves as a guideline for replicating predictive analytics across other conditions and EHR systems. In addition, this collaboration ensured the consistency of using real world data, reduced data processing errors, and maintained higher overall project quality and transferability. To this end, we shared the entire process—feature selection, model training, and all—in an open-source GitHub site along with clear instructions on how to utilize these models. We also created a user-friendly dashboard that shows ESRD risk in a simple way, giving researchers, doctors, and policy makers data they can utilize now. By freely sharing these tools, we lay the groundwork for future projects in predictive modeling and transfer learning.

### Role of Multimorbidity

Patients with diabetes frequently carry multiple co-existing chronic conditions—such as hypertension, cardiovascular disease, and obesity—that can interact additively or even synergistically to accelerate renal decline. By explicitly modeling these multimorbidity profiles through scalable joint and dynamic prediction techniques, our framework teases apart each condition’s contribution to end-stage renal disease risk, yielding more accurate, individualized forecasts. Incorporating multimorbidity is therefore essential both for precise risk stratification and for guiding tailored interventions to prevent or slow renal complications.

**MVP-CHAMPION 067: Long Covid**

**VA Lead: Shiuh-Wen Luoh**

**DOE Lead: Ravi Madduri**

### Rationale

Long Coronavirus 2019 disease (long COVID) is a chronic, disabling condition that affects an estimated 7% of adults and 44 to 48 million Americans^37^. Long COVID is projected to cost between $5,100 and $11,600 per case in the first year (primarily due to 92.5%–95.2% productivity losses). This could result in an annual societal cost of $2.01–$6.56 billion, including $1.99–$6.49 billion in productivity losses for employers and $21.0–$68.5 million in costs for third-party payers (based on a 6%–20% Long COVID probability)^37^. It is an incredibly heterogeneous disease with hundreds of symptoms. This type of manifestation has been observed for other infectious pneumonias, expanding the relevance of this project.

### Aim

to identify genetically-informed (or precision medicine) therapies, such as repurposing existing drugs to treat Long COVID.

### Data

MVP biobank provides 6,478 cases from multiple population groups. MVP linked to VA longitudinal clinical data Long COVID-19 is highly heterogeneous. We performed factor analysis to identify latent subtypes driving the diversity of symptoms. These included multiple subtypes: multi-organ (skin, heart, gastrointestinal), neurological (taste), Severe acute coronavirus 2019 disease, pulmonary, cardiometabolic, mental health, prevalent auto-immune disease, unknown, fatigue, joint pain which was accompanied by symptoms such as flushed skin, loss of taste, hypoxia, shortness of breath, hypertension, mood disorders, anaphylaxis, malaise and fatigue, joint pain and stiffness, cardiac arrhythmia, headache, pneumonia, chest CT, diabetes, depressive symptoms, persons with health hazards, chronic fatigue syndrome, osteoarthritis, localized, ulcer, viral symptoms, remdesivir, lipid metabolism disorder, substance use, neurological disorders, injection medication. Symptoms were measured in days noted (based on NLP) or recorded (based on ICD codes) & duration in months (last date – first date).

### Analysis

By comprehensive evaluation of the MVP clinical data, we were one of the two large genomic medicine databases, the other being HGI, in an advantaged position to comprehensively evaluate and develop subtypes of Long COVID using clinical notes. We are using the following three subtyping strategies: A data-driven machine learning algorithm, An expert curated algorithm based only on peer-reviewed studies, and A hybrid machine learning and curated algorithm.The metric for success is interpretability of the top 10 subtypes that explain >50% of variation in Long COVID symptoms. Machine learning explains more variation but is less interpretable, while expert curation has the opposite problem, so we are tuning a hybrid approach to balance predictive performance (>50% variation explained) and interpretability.

### Results

We identified two genetic variants for Long COVID among Veterans in MVP. The first variant was found in collaboration with the HGI and has led to 1 article under final stages of review at Nature Genetics^38^. We discovered another genetic variant for Long COVID in MVP Veterans of African ancestry, but not in other ancestry groups. We were able to find better agreement of long COVID subtypes and GWAS variants across ancestries. We developed subtypes of Long COVID using clinical notes and using these subtypes, We found greater associations in three subtypes among European and African ancestries: fatigue, pulmonary, and severe acute COVID-19 subtypes with the GWAS variants from the MVP. These subtypes were also validated independently in the variant from HGI.

### Efficiencies

The comprehensive clinical follow up information of the MVP uniquely positions us to continue to refine our subtypes of Long COVID to improve agreement across ancestries. Additionally, we discovered another genetic variant specific to MVP Veterans of African ancestry, not observed in other groups. This collaboration with HGI allowed us to leverage their established GWAS pipelines and shared statistical methods, enhancing the efficiency of our variant discovery efforts. In addition, the collaboration between the DOE scientists and domain experts at the VA allowed for the efficient use of algorithms within the Oak Ridge Leadership Computing Facility Knowledge Discovery Infrastructure (OLCF KDI) environment to run phenotyping algorithms, GWAS analysis, fine-mapping, and factor analysis to identify latent subtypes driving the diversity of symptoms. The contributions to others in the group include accessibility to run a high number of jobs in parallel within the OLCF KDI environment. In addition to the usage of the NLP algorithms to improve the ability to detect new cases of Long-COVID within the MVP biobank which will give higher power to perform the genetic analysis. This work would not have been possible without the collaboration between the Long-COVID genetic domain experts and computational scientists at the Argonne National Laboratory team who integrated several high-performance workflows for data analysis.

### Next Steps/Applications/Translation

We are refining our subtypes of Long COVID to improve agreement across ancestries. This includes adding diagnosis codes (ICD-10) and medications, e.g. adrenergic inhalants, to the clinical notes. Our work has focused on identification of the mechanisms of Long COVID, in particular identifying key genetic variants. For example, we found strong associations in fatigue, pulmonary, and severe acute COVID-19 subtypes with MVP-derived GWAS variants in both European and African ancestry groups. These subtypes were independently validated using the HGI-identified variant. Future steps include finding druggable genome applications. There are currently no approved therapy or even biomarkers for long COVID. The VA Causal group has created a druggable genome pipeline that allows us to identify genetic variant(s) and associate it with gene expression or protein level that specifically affect Long COVID^39^.The clinical translation of these findings is to provide essential clinical understanding of the mechanisms of different subtypes of long COVID to inform clinical research and management of long COVID.

Role of Multimorbidity: The complexity and variability of clinical presentations of long COVID presented a particularly challenging issue in the study of long COVID. Our study takes into account the temporal changes in symptom presentation, including the onset, frequency and severity of individual clinical features before, during the acute COVID infection and subsequent development of long COVID associated features. These efforts enable us to better define the subtypes of long COVID as well as understand the comorbidities that are associated with long COVID. Understanding these comorbidities presents an important opportunity to stratify our at risk Veteran populations for prevention and treatment intervention to mitigate long COVID.

**MVP-CHAMPION 068: Antipsychotic-induced Metabolic Adverse Effects**

**VA Lead: Ayman Fanous**

**DOE Lead: Kushbu Agarwal**

### Rationale

Mental health conditions including post-traumatic stress disorder, major depression, schizophrenia, and bipolar disorder affect 24% of the Veteran population. VA healthcare costs for these conditions exceed half a trillion dollars annually^40–42^. The advent of second generation antipsychotics (SGAs) has transformed mental healthcare,^43^ leading to improvements in the *symptoms* of illness, but they are also associated with significant metabolic adverse effects, including weight gain, hypercholesterolemia, and type II diabetes^44^, which increase risk of heart disease, stroke, kidney failure, and premature death. Furthermore, they lead many patients become non-adherent, leading to functional impairment, treatment resistance, and increased suicide risk. Precision medicine holds out the hope that an individual’s specific genetic makeup, lifestyle, and environment can be used to predict their response to treatment, including both beneficial and adverse effects^45^.

### Aim

We sought to develop artificial intelligence methods for predicting and reasoning about antipsychotic-induced metabolic adverse effects. Such models could accelerate the development of Precision Psychiatry modalities that can be used at the point-of-care to select treatments that maximize beneficial and minimize adverse effects. This could substantially reduce morbidity and mortality and reduce healthcare expenditures.

### Data

MVP and the VA EHR cohorts (690,000 patients on antipsychotics, the largest cohort ever studied for this purpose).

### Analysis

Using nine items from the Operationalised Criteria Computerised Diagnostic System representing psychotic, manic, depressive, and negative symptoms—aligned with our previously established four-factor model of schizophrenia^46^—we leveraged Large Language Models (LLMs) to extract clinically meaningful information. Our innovative methodology employed GPT-4 (achieving 0.75 concordance with physician raters), along with other LLMs on a sample of 24 Diagnostic and Statistical Manual 5c^47^ cases. Using our LLM-driven methodology and supercomputing capability at DoE, we processed more than 3 million unstructured clinical notes in order to extract patients’ functional impairment and crucial psychosocial factors including sleep patterns, exercise habits, and family structure. The psycho-social factors, primary comorbidities, medications and lab results were then integrated in the ML pipeline to identify the protective and risk inducing factors for antipsychotic induced adverse effects.

### Results

This project has yielded a number of potentially impactful findings. We examined the association of increases in body mass index after six months of antipsychotic treatment with genetic and demographic factors and comorbidities in a cohort of 137,771 genotyped participants. Latino ancestry and type 2 Diabetes Mellitus PRS were positively correlated, while SGA use and Bipolar Disease PRS were inversely correlated. Several genome-wide significant loci were identified, most notably, in the gene *PRDM16*, which determines brown adipocyte cell fate and is considered a potential therapeutic target for obesity^48^. We discovered that marital status serves as a significant protective factor against antipsychotic-induced weight gain, which remained robust when controlling for initial metabolic health, functional impairment, and medication dosage. Our LLM work could inform that of other teams who are also interested in extracting meaningful variables from free text. Furthermore, the social determinants of health and functional impairment variables we extracted could be used by researchers to study other outcomes, such as diabetes and heart failure.

### Efficiencies

this project brings together the clinical and genomics expertise of Drs. Ayman Fanous and Silviu-Alin Bacanu of the Phoenix VA, with the AI and analytic expertise of Drs. Khushbu Agarwal, Sutanay Choudhury, and Ben McMahon of the DOE. It could only have been implemented by this team, as they have access to both the VA data as well as *Frontier*, the world’s first exascale supercomputer, for analyses. The analysis of this dataset could not be performed otherwise, on account of its sheer size and the computational intensiveness of the cutting-edge analytic procedures required. Furthermore, we were able to benefit greatly from the experience of other ongoing VA-DOE projects, such as Dr. McMahon’s work in suicidality, which optimized study design.

### Next Steps/Applications/Impact

The unprecedented scale and novel methodology of this work deliver findings with immediate clinical applications. By developing methods capable of extracting complex clinical phenotypes from electronic health records, we have created a framework that enables: a) identification of new druggable targets related to brown adipocyte development that industry partners could use as the basis for novel therapeutics; b) risk stratification using PRSs, which can enable more rational treatment planning; c) clinical subtyping of patients from massive EHR datasets for downstream prediction models; and d) determination of other pharmacogenomic phenotypes, particularly symptom improvement, which could revolutionize models of differential treatment response. The generalizability of these findings across such a large and diverse patient population positions this work to significantly advance personalized approaches to psychiatric care while addressing one of its most insidious adverse effects.As such, our work could fit into several phases of the translational research continuum. The identification of *PRDM16* could suggest potential transgenic animal model studies to more specifically elucidate the impact of genetic variation on functions of this gene (T0). PRSs could be studied as prognostic factors in clinical trials (T2). More routine assessments of individual patients’ social relationships by their providers could help identify those at higher risk of metabolic adverse effects (T3). Finally, promoting greater levels of social support for individuals with serious mental illness could have improved health outcomes for this vulnerable segment of society (T4).

### Role of Multimorbidity

We were able to visualize the complex interplay of multiple modalities, such as medication use, comorbidities, and substance misuse in the prediction of adverse events, using existing figures and graphs.

